# DEVELOPMENT AND CONTENT VALIDITY OF THE HANDWASHING INDEX (HWI)

**DOI:** 10.1101/2024.07.20.24310756

**Authors:** Emmanuel Lamptey, Ephraim Kumi Senkyire, Doris Selorm Agoha

**Affiliations:** KAAF University college, Ghana; Ga-West Municipal Hospital, Ghana Health Service

**Keywords:** content validity, instrument development, hand hygiene, population

## Abstract

Handwashing is the simplest way to stop the spread of infections and maintain health. To the best of our knowledge, there is no validated scale to measure this habit. Given the need to raise awareness and its importance, a study was undertaken to develop, test, and validate the HWI. The Physical Activity Questionnaire for Adolescents (PAQ-A) was adapted and substantially modified to draft the first version of the index (HWQ). The COnsensus-based Standards for the Selection of Health Measurement Instrument (COSMIN) guided our approach. The HWQ The questionnaire consisted of six questions with a five-point scale response for each item, ranging from the lowest [1] to the highest activity [5] as scoring criteria. We followed a four-stage procedure in validating the instrument: Stage I: items were created from the review of the relevant literature (PAQ-A, COSMIN); Stage II: the study targeted and recruited 57 health professionals from different backgrounds and vetted them as decliners, non-regulars, and regular hand washers through a scoring process.

This crucial phase ensured that the respondents had hands-on experience in handwashing in addition to their expertise. In Stage III, 10 respondents from the regular hand washers were randomly selected as expert judges. They were divided into two (2) groups (Group one and Group two) of four and six members, respectively. Further comprehensibility and comprehensiveness of the HWQ were established through a review by the four members of Group 1, and revisions were made to create HWQ-I. At stage IV, the HWQ-I was administered to the six members of the second group for ratings and content validity index calculations. The final draft of the HWI includes 4 items. Item CVI ranges between 0.8 and 1.The average scale CVI was 0.9.

The HWI demonstrated excellent content validity, showing its relevance for monitoring handwashing, and individuals can track their compliance, promoting a healthy life. We propose further studies to test the psychometric properties of the scale.

## Introduction

Handwashing is an effective way of keeping diseases at bay, and the emergence of the COVID-19 pandemic reinforced its importance, which has received considerable attention [1] [2] [3] [4]. COVID-19 spreads through direct and indirect sources such as contact with contaminated objects, sneezing, coughing, and secretions from the nose and mouth. Handwashing is critical for interrupting this chain and is the main cornerstone of prevention [2].

According to a study conducted in July 2020, the incidence of coronavirus cases and related deaths declined in 6064 adults who adhered to the WHO handwashing guidelines [5] [6].

In other studies, handwashing has proven effective against the acquisition of SAR-COV-2 and other related viruses such as SARS-CoV-1 and influenza [7]. Proper handwashing prevents the dissemination of SARS-CoV-2, eventually killing the enveloped virus [4]. It controls the propagation of the virus more than alcohol-based sanitizer for hand decontamination [4].

Handwashing was a step taken to keep people, workers, and families safe during the outbreak [8].

When done properly, it is associated with a 30-48%, 23%, 50%, and 27% reduction in the risk of endemic diarrhea, acute respiratory infections, pneumonia, and related infant deaths, respectively [9]. The propagation of other equally infectious diseases such as Ebola, hepatitis, cholera, and shigellosis can be stopped with this practice [9]. Despite its significance as the most cost-effective public health intervention, non-adherence is alarmingly widespread and compliance is still very low, although it is a simple behavior [10]. Multiple studies have reported varying hand hygiene habits in the pre-and post-COVID-19 era. A study, conducted in 2015, estimated that only 26.2% of respondents who visited places of convenience with potential fecal contact, washed their hands [11]. Another study in Denmark between the years 2020 and 2022 showed changing hand hygiene habits, with Scandinavians slipping back into their old ways when the COVID-19 pandemic subsided [12]. In a related development, compliance among healthcare workers fell back to pre-pandemic levels (51% vs 90%) [13].This study also projected that the public is likely to follow the influence of the medical community [13] [14]. In summary, people tend not to worry much about handwashing anymore because COVID-19 is no longer a threat [12] [15]. Although this can be partially attributed to the lack of flowing water and soaps in deprived regions of the globe, this behavior has declined in many resourced countries [11] [16]. Handwashing has been a lifesaving intervention in the history of humankind, prolonging the lifespan of individuals to approximately 80 years [17]. A country’s handwashing culture can predict the degree of infection spread [18]. Therefore, washing hands is the right thing to do or the pathway to improved health, irrespective of any pandemic [19]. To promote such behavior, the public must not only embrace and normalize it as the pillar of infection prevention but also mandate it at critical times (before eating and after defecation) and when someone comes home from public domains [3].

The theoretical implication is that everyone should adopt this behavior to improve their daily health [19]. There are no definitive reports of any tools used to measure handwashing in individuals. A new instrument is needed to measure this behavior in terms of making healthy lifestyle choices and habits. Evaluation of handwashing practices is a vital strategy for improving hand hygiene, making it possible to measure changes induced by its implementation [21]. This study aimed to develop the Handwashing Index (HWI) and test its content validity.

## Methods

We developed the handwashing index and its content validation in four stages according to the COSMIN guidelines. The research team, which includes researchers who are well-grounded in nursing, arrived at a consensus to undergo the following four stages.

### Stage 1: Development of the Handwashing Questionnaire (HWQ)

The authors first developed a tool that measures handwashing and enhances its content validity. The tool was adapted from the Physical Activity Questionnaire for Adolescents (PAQ-A). The PAQ-A was developed by Kowalski et al. [22]. The purpose of the PAQ-A is to assess physical activity and self-reported measures in adolescents.

The PAQ-A was substantially modified, changing the content of items to fit the context of handwashing and monitoring the effective participation of this behavior. Items unrelated to the construct and judged not applicable were removed from the instrument. Relevant literature on quantitative evaluation and questionnaire development were reviewed to ensure that each item on the tool focuses on activities that individuals can perform to maintain a healthy life and well-being [23]. The above sources were used to create the initial draft of the instrument, which consists of six items (Appendix A-HWQ). A continuous option scale that starts from the lowest to highest handwashing activity was selected to answer the items in the instrument. The lowest and highest responses were 1 and 5, respectively.

### Stage II: Recruitment of the target population and panel of experts

In the second phase, we aimed to identify the target population (professionals from all health disciplines) and use appropriate methods to justify their recruitment as a panel of experts. Participants were recruited via email from professional groups, networks, newsletters, and Facebook conference pages in Ghana. The study was also advertised in circular emails and on the social media handles of professional organizations. Participants were eligible if they were registered health professionals working in a clinic, hospital, health organization, or training institution (university) and had consented to participate. Participants were asked to indicate this before a link to the survey was redirected. We recruited participants from September 18 to November 4, 2023, with a data collection period lasting 4 weeks.

The first draft questionnaire (HWQ) was completed by 57 health professionals working in different fields. Convenience sampling was employed to obtain data from these accessible populations because this study considered their readiness to make time and respond.

Socio-demographic and professional data (years of work experience, professional background, gender and qualification) from these respondents were also obtained. Respondents who completed the questionnaire were screened and scored.

Handwashing entails conscious effort, and to ensure this, we presented a weighting method to screen the target population and judiciously select experts who are consistent in this practice by scoring the questionnaire (Appendix A-HWQ) [24]. We argued that the target population should be scored and categorized to define them as expert judges. Thus, those who perform handwashing as a daily habit are equipped with the necessary facilities.

The aim was also to identify the varying degrees of performance among the target population. The lowest response was recorded as one (1), whereas the highest response was five (5). The mean of the 6 items (1, 2, 3, 4, 5, and 6) was taken to give the average score for each participant. Mean scores of 0–1, 2–3, and 4–5 were categorized as decliners, non-regulars, and regular hand washers, respectively. At this stage, the participants did not assess or comment on the relevance, comprehensiveness, or comprehensibility of the questionnaire and were not given that option.

We excluded participants who provided incomplete feedback on the questionnaire. Participants who obtained average scores of 4-5 after scoring the questionnaire were classified as expert judges out of the target population and were focused on for the next stage (Stage III) of the study.

### Stage III: Development of the Outcome Measurement Instrument

In the third stage, the research team randomly selected 10 people who were categorized as regular hand washers or expert judges. This set scored an average of 4-5. Lynn (1986) advised a minimum of three and a maximum of ten experts; hence, more than 10 experts are unnecessary. For substantial item improvement, we initiated two rounds of expert review during this phase. The 10 expert judges were divided into two (2) groups of four (Group-1) and six (Group-2). Group 1 was given clear instructions to comment and provide guidance in revising, deleting, or substituting items of the HWQ. They were not asked to rate the relevance or provide I-CVI values.

The following comments were made. Item 4 of the questionnaire (Appendix A-HWQ) causes the variable. Handwashing may be caused by an activity or work. One does not need to engage in an activity that requires hand hygiene; however, it should be self-care. Remembering the frequency of handwashing within the last 7 days is difficult because of busy schedules and lifestyles. People do not wash their hands in the evening because of their normal bathing routine. Revisions were made on the basis of the above comments, which created the second draft (Appendix B-HWQ-I) with 5 items. The number of days was reduced to three, Item 3 was deleted, and item 4 rephrased not to be a cause of the variable. Then, all the items were reviewed by another expert proficient in the English language to make them clear and simple to understand, thus increasing comprehensibility.

### Stage IV: Content Validation Process

A content validation form C together with HWQ-I was then administered to the six members of Group 2. They were asked to rate each scale item in terms of its relevance to handwashing. Rating was done on a 4-point ordinal scale to avoid a neutral midpoint as follows: 2=somewhat relevant, 1=not relevant, 4=highly relevant, 3=quite relevant. Scores of 1 and 2 are irrelevant, whereas 3 and 4 are relevant.

**Figure 1:**
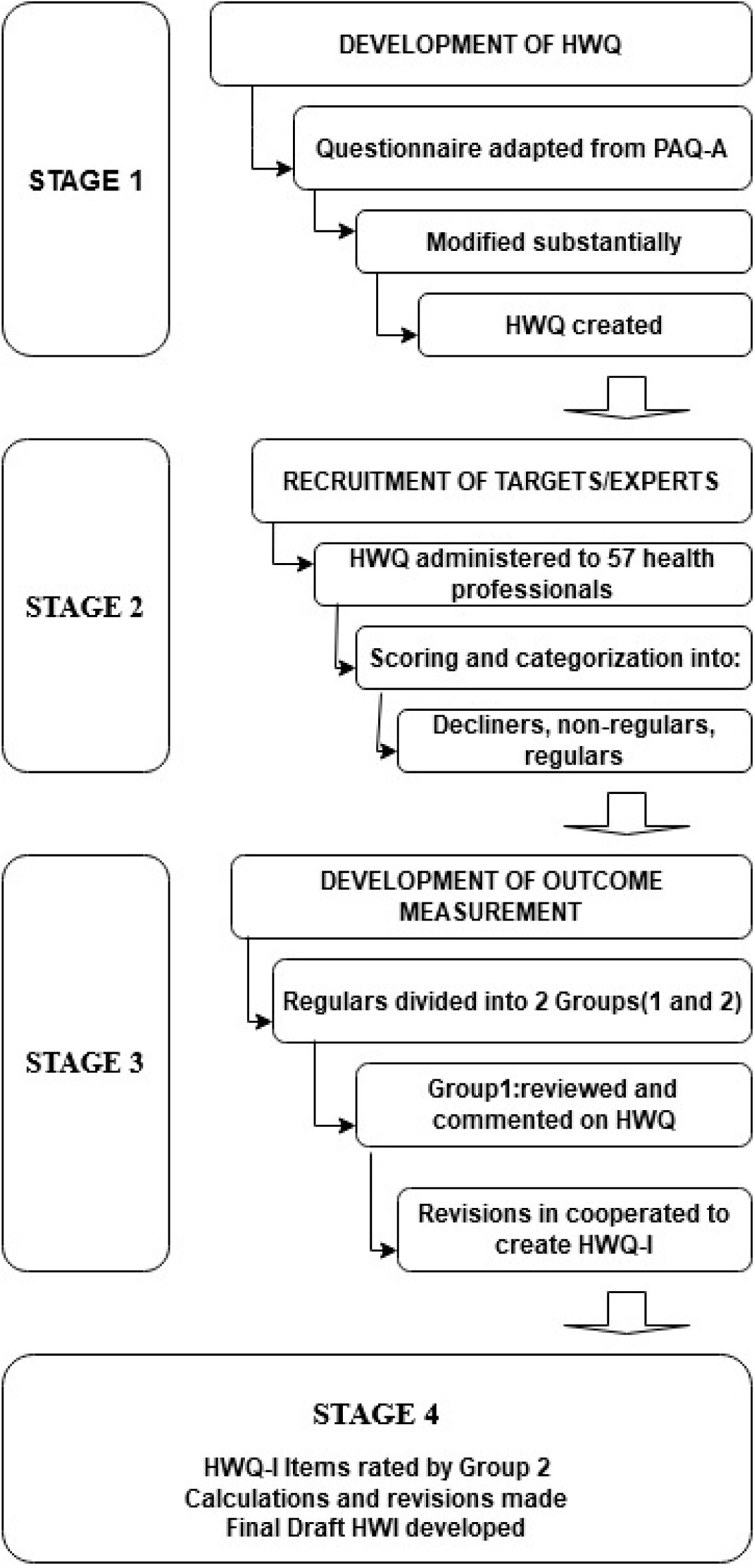
Study stages

## Ethical Considerations

The study followed the instructions and protocol requirements of the Ghana Health Service Review Committee (GHS-ERC-045/08/23). We informed the respondents that participation was not mandatory. The purpose of the study was explained as follows: the findings will not be used beyond this purpose, nor will they be an evaluation or judgment of the value of their practice. Participants were free to withdraw at any time, and confidentiality was fully maintained with no participant identifiers.

## Data Analysis

We analyzed the professional and socio-demographic characteristics of the participants at Stage II using descriptive statistics and subsequently their scoring and categorization as decliners, non-regular, and regular hand washers.

The ratings from six expert judges at Stage IV were dichotomized into two (2) categories: scores one (1) and two (2) were irrelevant, and scores three (3) and four (4) indicated relevant items. We computed experts in agreement as the number of experts who agreed relevant to the items, e.g., those who rated the item as 3 or 4.

Item Content Validity (I-CVI) was calculated by dividing the experts in agreement by the number of experts. Universal Agreement (UA) was obtained by assigning a score of 1 to the item that achieved 100% expert in agreement and 0 to the item that did not achieve 100% expert in agreement. The HWI content validity (S-CVI/Ave) was computed by averaging the I-CVI score across all items. Proportion relevant (PR) is the number of items agreed upon by the judges as relevant and given a score of 1 and a score of 0 if not relevant.

The socio-demographic characteristics of the target population at the second stage are presented in Table 1. A total of 57 health professionals responded as the target population, the majority being females (n=37), having a first degree as their highest qualification (35.1%) with an average of 11.5 years of work experience.

**Table 1:** Socio-demographic data of the target population engaged at stage 1 (N=57)

| **Variable** | **N** | **Mean ± SD** |
| --- | --- | --- |
| **Age** |  | 38.39±12.38 |
| **Gender** |  |  |
| Female | 37(65%) |  |
| Male | 20(35%) |  |
| **Area of Practice** |  |  |
| Public Health nursing | 8(14%) |  |
| Community health nursing | 3(5.3%) |  |
| General nursing | 27(47.4%) |  |
| Health administration | 1(1.8%) |  |
| Internal medicine | 9(15.8%) |  |
| Midwifery | 4(7%) |  |
| Ophthalmology | 1(1.8%) |  |
| Pediatrics | 1(1.8%) |  |
| Obstetrics | 3(5.3%) |  |
| **Highest Qualification attained** |  |  |
| Certificate | 17(29.8%) |  |
| Diploma | 19(33.3%) |  |
| Degree | 20(35.1%) |  |
| Masters | 1(1.8%) |  |
| **Years of Experience** |  | 11.54±8.74 |
| HDWI-Hand washing index, SD=Standard deviation |  |  |

The categories after scoring are presented in Table 2. We recorded one decliner, 25 regulars, and 31 non-regular hand washers (Table 2). The content validity ratings from six expert judges (categorized as regulars) are presented in Table 3. We found that the I-CVI (HWQ-I) for the 5 items ranged from 0.5 to 1.0 (Table 3). Item 3* fell below the acceptable cutoff of of 0.78 [25], the remaining 4 items scored 0.78, and the S-CI (average) and S-CVI(UA) values were 0.82 and 0.9, respectively. Therefore, we made the following modifications: Item 3 was deleted from HWQ-I to give the final draft (HWI) of the scale. After implementing the above-suggested change, I-CVI and S-CVI were recalculated for the second time. S-CVI achieved an acceptable value of 0.9[26]. The mean I-CVI values for calculating the content validation resulted in an instrument with 4 items. After this, no item achieved an I-CVI value below 0.8 and the S-CVI was 0.9. Therefore, no further computations, revisions, or deletions were necessary.

**Table 2:**
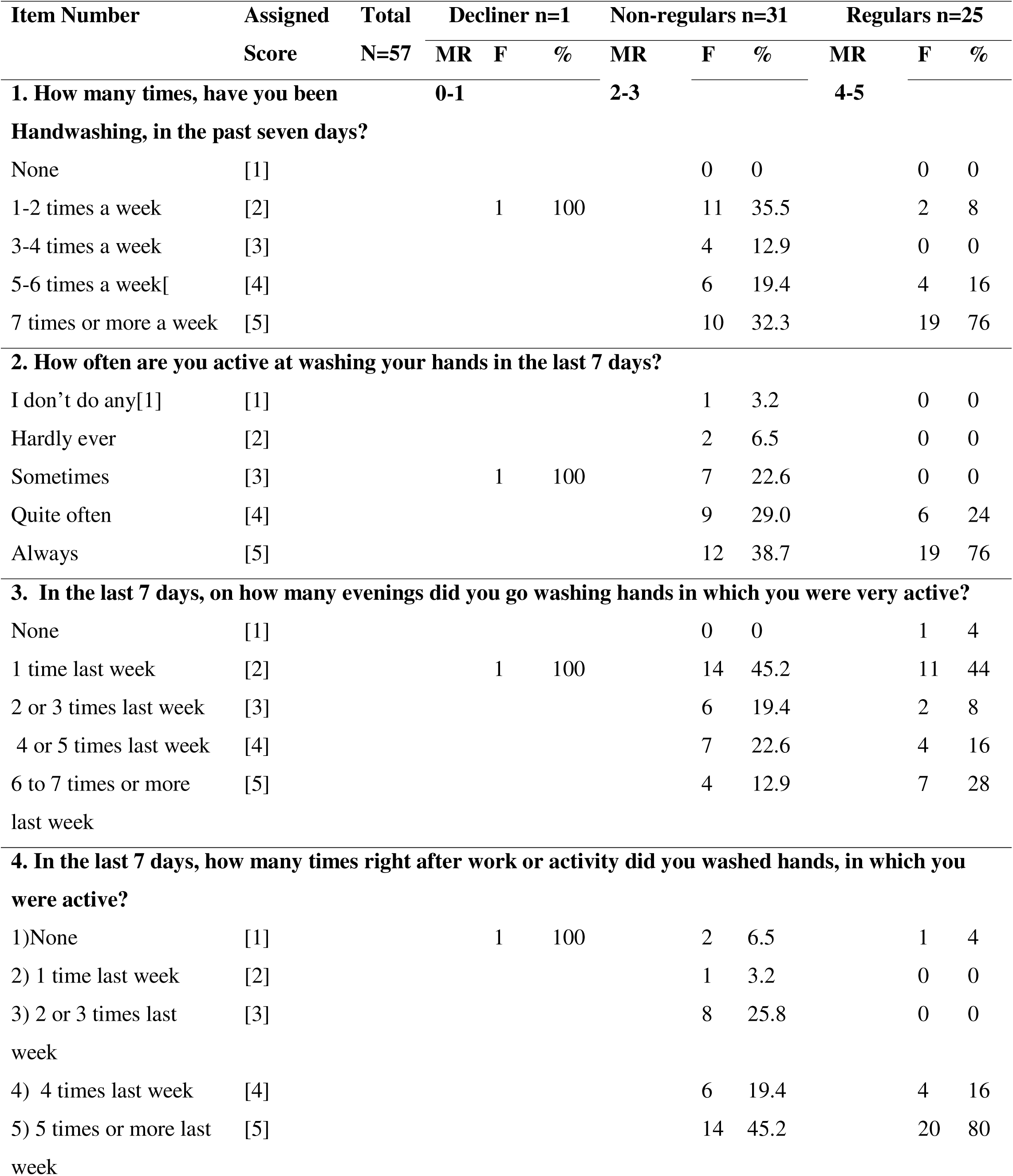
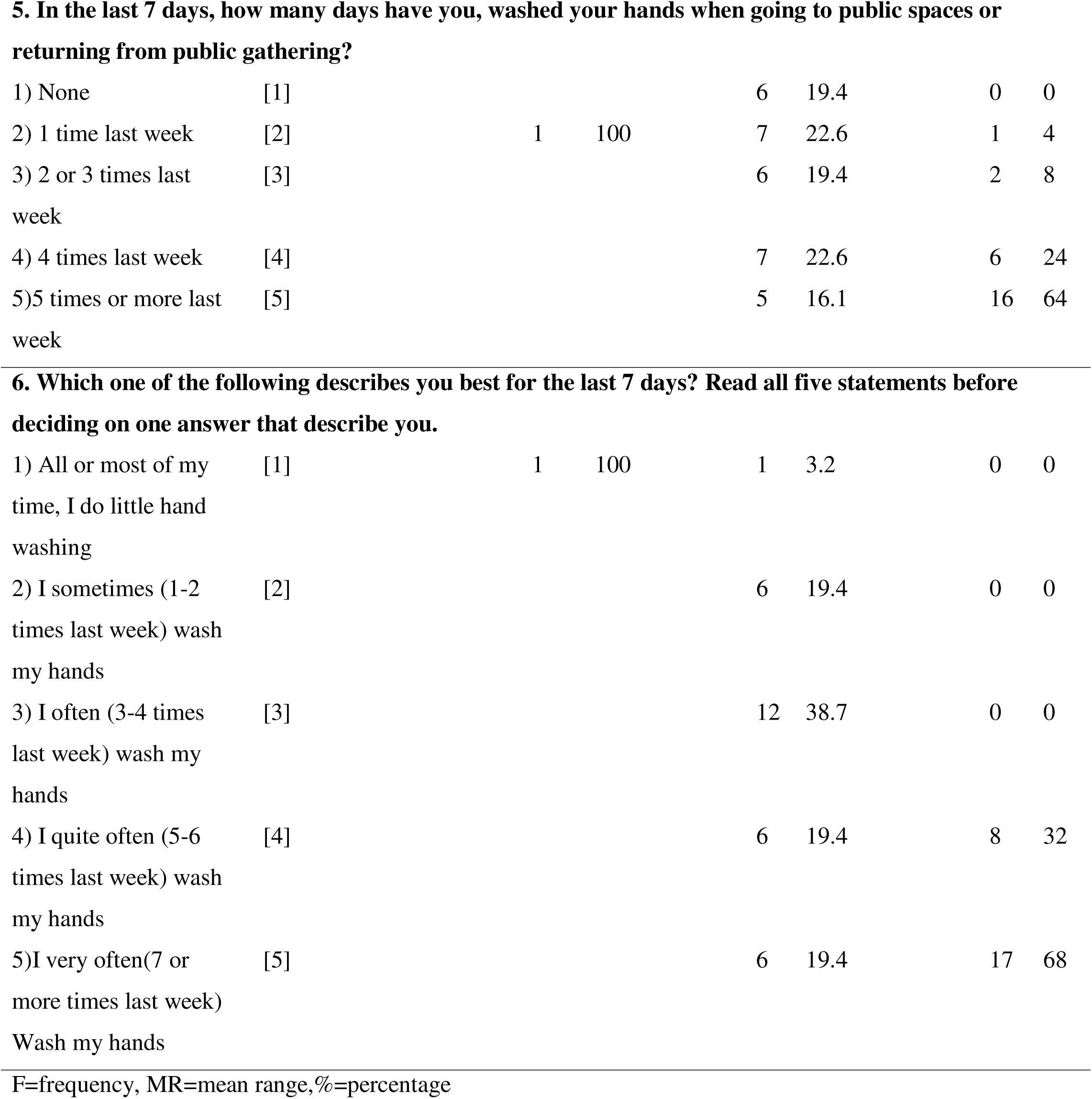
Scoring of Participants who responded as target population at Stage 1.

**Table 3:** Ratings of the 5-items of HWQ-I by the six experts at stage 4 on a 4-Point relevance scale.

| Item No. | Expert 1 | Expert 2 | Expert 3 | Expert 4 | Expert 5 | Expert 6 | Expert in Agreement | I-CVI | UA |
| --- | --- | --- | --- | --- | --- | --- | --- | --- | --- |
| 1 | 4 | 3 | 3 | 3 | 3 | 4 | 6 | 1.0 | 1 |
| 2 | 4 | 2 | 3 | 3 | 4 | 4 | 5 | 0.8 | 0 |
| 3* | 2 | 1 | 2 | 3 | 4 | 3 | 3 | 0.5 | 0 |
| 4 | 4 | 3 | 3 | 3 | 4 | 4 | 6 | 1.0 | 1 |
| 5 | 4 | 2 | 3 | 3 | 4 | 4 | 5 | 0.8 | 0 |
| PR | 0.8 | 0.4 | 0.8 | 1 | 1 | 1 |  | S-CVI | S-CVI |
| Ave= |  |  |  |  |  |  |  | Ave= | UA=0.4 |
| 0.83 |  |  |  |  |  |  |  | 0.82 |  |

I-CVI, item content validity index
S-CVI/UA-Scale level content validity, Universal agreement.
PR: Proportion relevant

**Table 4:**
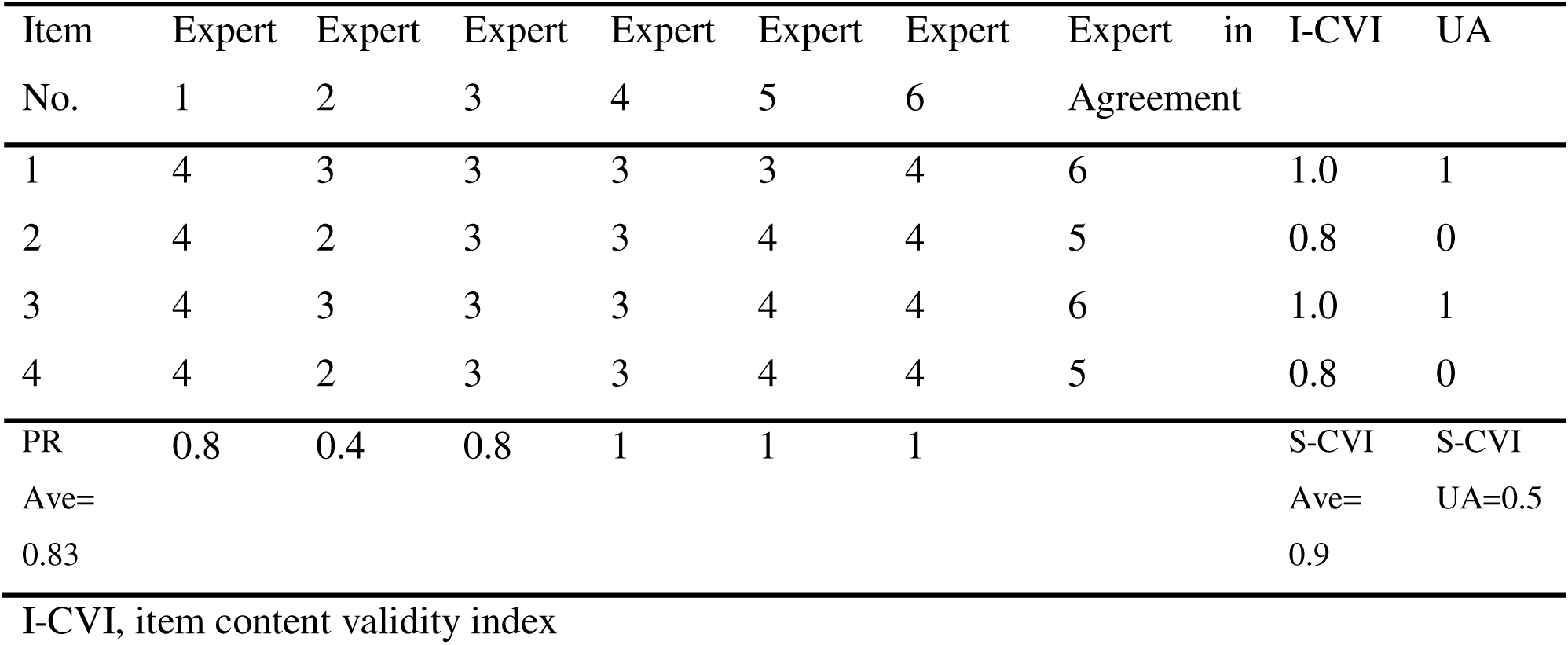
Recalculations of ratings of the 4-items of the HWI.

| Item No. | Expert 1 | Expert 2 | Expert 3 | Expert 4 | Expert 5 | Expert 6 | Expert in Agreement | I-CVI | UA |
| --- | --- | --- | --- | --- | --- | --- | --- | --- | --- |
| 1 | 4 | 3 | 3 | 3 | 3 | 4 | 6 | 1.0 | 1 |
| 2 | 4 | 2 | 3 | 3 | 4 | 4 | 5 | 0.8 | 0 |
| 3 | 4 | 3 | 3 | 3 | 4 | 4 | 6 | 1.0 | 1 |
| 4 | 4 | 2 | 3 | 3 | 4 | 4 | 5 | 0.8 | 0 |
| PR | 0.8 | 0.4 | 0.8 | 1 | 1 | 1 |  | S-CVI | S-CVI |
| Ave= |  |  |  |  |  |  |  | Ave= | UA=0.5 |
| 0.83 |  |  |  |  |  |  |  | 0.9 |  |

I-CVI, item content validity index

## DISCUSSION

Considering the need to stop the spread of infections and the importance of handwashing to achieve this task. We developed a handwashing index (HWI) for the general population and tested its validity. To the best of our knowledge, the HWI is the first instrument to measure handwashing and comprehensively monitor this behavior. In the current study, a representative sample (regulars, n=25) from which we derived the evaluations at stage IV was solicited, described and justified as expert judges who practiced this particular behavior as a daily routine aside from health expertise.

They were screened out of the target population of judges (n=57) as those who had extensively practiced handwashing and had the procedural knowledge, skills, and necessary facilities to execute the function. The scoring process at stage II ensured that the participants had attained proficiency and experience in handwashing. The remaining target population who were categorized by as Decliner(s), n=1, non-regulars, n=31 suggest limited access to handwashing facilities and were less likely to wash their hands. These observations indicate that there is no reliable proxy for measuring handwashing by asking participants to report their behavior.

In line with DeVellis (2022), non-regulars and decliners were target population judges, whereas regulars were expert judges because of their procedural knowledge and experience [27] [28] [29]. Although the opinion review of the target population is essential and part of the basis for content validity [29] [30] [31], we used the regulars because of limited resources [31].

We computed two types of CVIs: the content validity of individual items (I-CVI) and the content validity of the overall scale(S-CVI) and reported them according to the data collected from expert judges. Lynn (1986) advocated I-CVI in the vicinity of 0.8 when there are six or more experts [25].

For the data in Table 3, all items met Lynn’s criteria or were within the acceptable range, except item 3*[25]. Item 3 had an I-CVI of 0.5, meaning it was inadequate, and the remaining responses scale obtained an I-CVI ≥ 0.8. A total of the 5 items obtained a CVI average (S-CVI) of 0.82, which implies that 82% of the items achieved a very relevant rating (3 or 4) by all the expert judges. However, an S-CVI (average) of 0.82 is acceptable only in cases where there are two raters.

In situations where there are more than two judges, Lynn explicitly recommended 0.9 and the SCI-UA extension. In addition, CVI can be a chance factor, and to incorporate a standard that eliminates this error, Lynn recommended that I-CVI values should not be lower than 0.78.

According to Polit and Beck, such I-CVI information can be used in revising, deleting, or substituting items on the scale; therefore, item 3 was discarded to give the final scale (HWI) [32].

Revisions are permissible [33].Flight et al. (2011) developed an initial item pool of 122 and finished the scale with only 43 items [33]. Several studies have also reported a reduction of more than 50% [34]. In this study, the number of items was only reduced by 10% because item 3 was excluded due to inadequacy.

Following universal congruent rating by all the raters (SCI-UA 0.4), only two items (Items 1, 4) out of the five items received a relevant rating (3 or 4). S-CVI/Ave is the same as the average congruence percentage (ACP), according to Waltz et al. (2005), and should be 0.9 not 0.8 as the standard criterion for accepting S-CVI [35].

The SCI/Ave method is preferred for scale level than S-CVI/UA because a universal agreement is stringent where there are six 6 experts in the validation panel and a biased viewpoint is avoided, but we reported both [35].

Compared with other instruments, HWI can measure the frequency of hand hygiene and the general practice of habit in both resourceful and resource-strained settings. HWI has multifaceted advantages for the population. It does not look at the WHO’s five moments (before patient contact, after body fluid exposure, before an aseptic task, after patient contact, and after contact with patients surrounding) restricted only to health workers. It can be used anywhere without sophisticated machines, power systems, or technology, and the issues of cost, equipment, location, and personnel requirements cannot render it inappropriate.

The tendency of behavior modification under direct observation can be minimized; therefore, its validity is not subject to the Hawthorne effect because the general population can self-administer at their convenience. Difficulties in asking or observing people to wash their hands and their subsequent biases can be avoided. Hence, more truthful or valid responses are expected.

HWI can identify adherence to handwashing according to the scoring scheme. If the respondent scores higher on the index, they are evaluated as regular hand washers. Because this scale is novel, it can be used as a standard for assessing hand hygiene performance and for educating and engaging health workers and the public in hygienic campaigns. Thus, HWI can be considered an innovative tool that will allow infection perfectionists to track the success of their intervention, thus reducing the incidence of acquired infection.

The HWI not only measures compliance with handwashing, but also how active individuals practice it, which enhances access to health.

This will contribute to increasing hand hygiene commitment and actions to achieve global targets. The public can receive individualized feedback on their frequency of handwashing because there is now a tool to monitor this practice. Raising awareness and priority and bringing about measurable improvements in compliance.

There is literature, policies in workplaces, posters, and graphical messages reminding people about the importance of handwashing for health, and it is endorsed by governments, schools, civil society organizations, NGOs, and private companies. Measurement is crucial for complete success. Individuals can know for themselves if they are properly performing this task to keep safe and their commitment to meeting regulatory standards of ensuring cleanliness and safety. This will foster a sense of responsibility and accountability as they can track their hand hygiene practices. Based on the HWI scores, areas for improvement, lifestyle modification, training, and reinforcing protocols can be targeted.

In addition, HWI offers the ability to assess the history of handwashing upon returning from a public gathering. Public gatherings cannot stop, and to save millions of lives, it is essential to consider this aspect in an evaluation tool. Therefore, evaluating the habits of individuals is necessary to define characteristics and focus on specialized investigation.

### Strength of the study

The study began with a strong conceptualization of the construct developed from the PAQ-A. We screened and judiciously selected experts with clear instructions. Two phases of expert review were conducted for substantial item improvement (Stage III and IV). The assessment of comprehensibility by experts provided an important contribution by anticipating situations that would make understanding of the scale difficult. Detailed information on the calculation methods was clarified in the content validation process. We have reported the I-CVI values for the items in each phase.

### Limitations

This study has geographical limitations. We adopted a convenient sampling method to recruit experts from Ghana. However, these experts were from diverse medical backgrounds, thus minimizing bias. We recommend future studies with international experts or experts from many countries to further validate the index.

### Implications for Nursing Practice

The HWI proved to have good content validity. It is available for future validation processes. When this is done, it will have a significant impact on nursing care and patient safety. The possibility of self-reported responses or assessment associated with the scale will remind nurses of their adherence to handwashing and the need to be regular handwashers. This will help maintain standards in the consistent practices of the habit. It will impact their knowledge, self-management, and compliance with infection control practices, translating into positive actions and attitudes.

## Conclusion

In this study, we developed a new instrument that measures handwashing and tested its validity with positive results. Because there is no such tool to evaluate this in individuals, HWI can be a valuable and essential instrument for self-education, clinical practice, and community protection. It can be used to tailor interventions aimed at improving handwashing in the global community.

## Data Availability

All data produced in the present study are available upon reasonable request to the authors

## Acknowledgment

The authors acknowledge the efforts of all experts who accepted to participate in the study

## Acknowledgment

None

## APPENDIXES

### APPENDIX A-THE HANDWASHING QUESTIONNAIRE (HWQ)

1. How many times, have you been hand washing, in the past seven (7) days?

1. None
2. 1-2 times a week
3. 3-4 times a week
4. 5-6 times a week
5. 7 times or more a week
2. How often are you active at washing your hands in the last 7 days?

1. I don’t do any
2. Hardly ever
3. Sometimes
4. Quite often
5. Always
3. In the last 7 days, on how many evenings did you go washing hands in which you were very active?

1. None
2. 1 times last week
3. 2 or 3 times last week
4. 4 or 5 times last week
5. 6 to 7 times or more last week
4. In the last 7 days, how many times right after work or activity did you washed hands, in which you were active?

1. None
2. 1 time last week
3. 2 or 3 times last week
4. 4 times last week
5. 5 times last week or more
5. In the last 7 days, how many times have you, washed your hands when going to public spaces or returning from public gathering?

1. None
2. 1 time last week
3. 2 or 3 times last week
4. 4 times last week
5. 5 times last week or more
6. Which one of the following describes you best for the last 7 days? Read all five statements before deciding on one answer that describe you.

1. All or most of my time, I do little hand washing
2. I sometimes (1-2 times last week) wash my hands
3. I often (3-4 times last week) wash my hands
4. I quite often (5-6 times last week) wash my hands
5. I very often (7 or more times last week) washed my hands

[Number of Items-6]

### APPENDIX B-HWQ-I

**HWQ-1 (**Questionnaire after first round of expert review at Stage III)

1. How many times, have you been hand washing, in the past **three (3)** days?

1. None
2. 1-2 times
3. 3-4 times
4. 5-6 times
5. 7 times or more
2. How often are you active at washing your hands in the last **3 days**?

1. I don’t do any
2. Hardly ever
3. Sometimes
4. Quite often
5. Always
3. In the last three days, how many times have you deliberately washed your hands without getting involved in any activity?

1. None
2. 1 time
3. 2 or 3 times
4. 4 times
5. 5 times or more
4. In the last 3 days, how many days have you, washed your hands when going to public spaces or returning from public gathering

1. None
2. 1 time
3. 2 or 3 times
4. 4 times
5. 5 times or more
5. Which one of the following describes you best for the last 3 days? Read all five statements before deciding on one answer that describe you.

1. All or most of my time, I do little hand washing
2. I sometimes (1-2 times) wash my hands
3. I often (3-4 times) wash my hands
4. I quite often (5-6 times) wash my hands
5. I very often (7 or more times) washed my hands

**[Number of Items-5]**

### VALIDATION FORM C

Dear Expert/Participants

I am conducting a research about **HANDWASHING (HW)** in form of survey. This survey contain **5 ITEMS (QUESTIONS)**. I need your judgment on the **DEGREE OF RELEVANCE OF EACH ITEM** to the measured Concept **(HW).** The instructions are provided below to guide you in your analysis. Please use the rating scale and rate each item as objective as possible in your own views.

1: Not Relevant

2: Somewhat Relevant

3: Quite Relevant

4: Highly Relevant

#### INSTRUCTION

This instrument is for the expert evaluator to assess the relevance of the Instrument you are validating. Indicate your judgment of relevance to each ITEM by ticking the approach box.

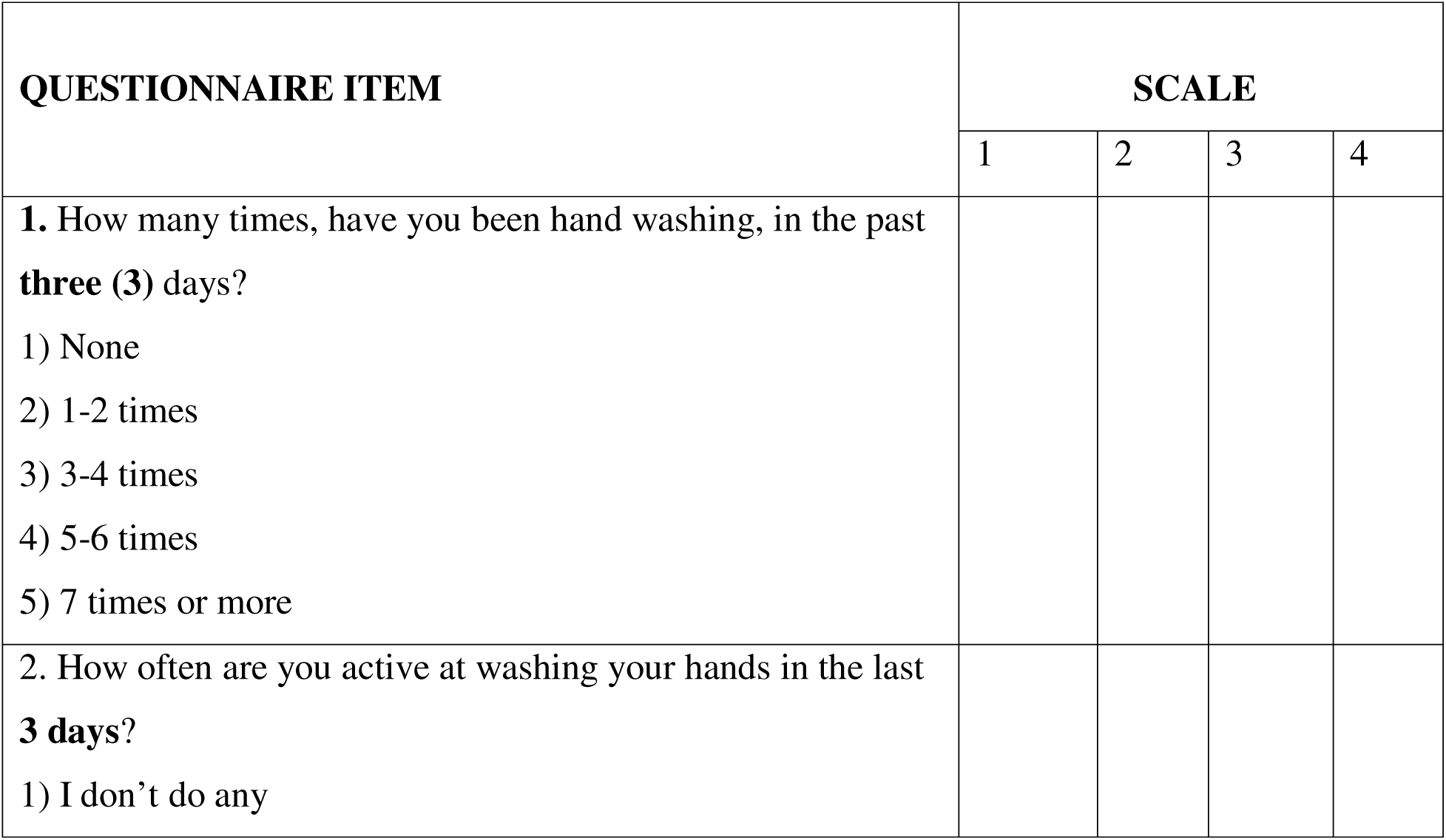
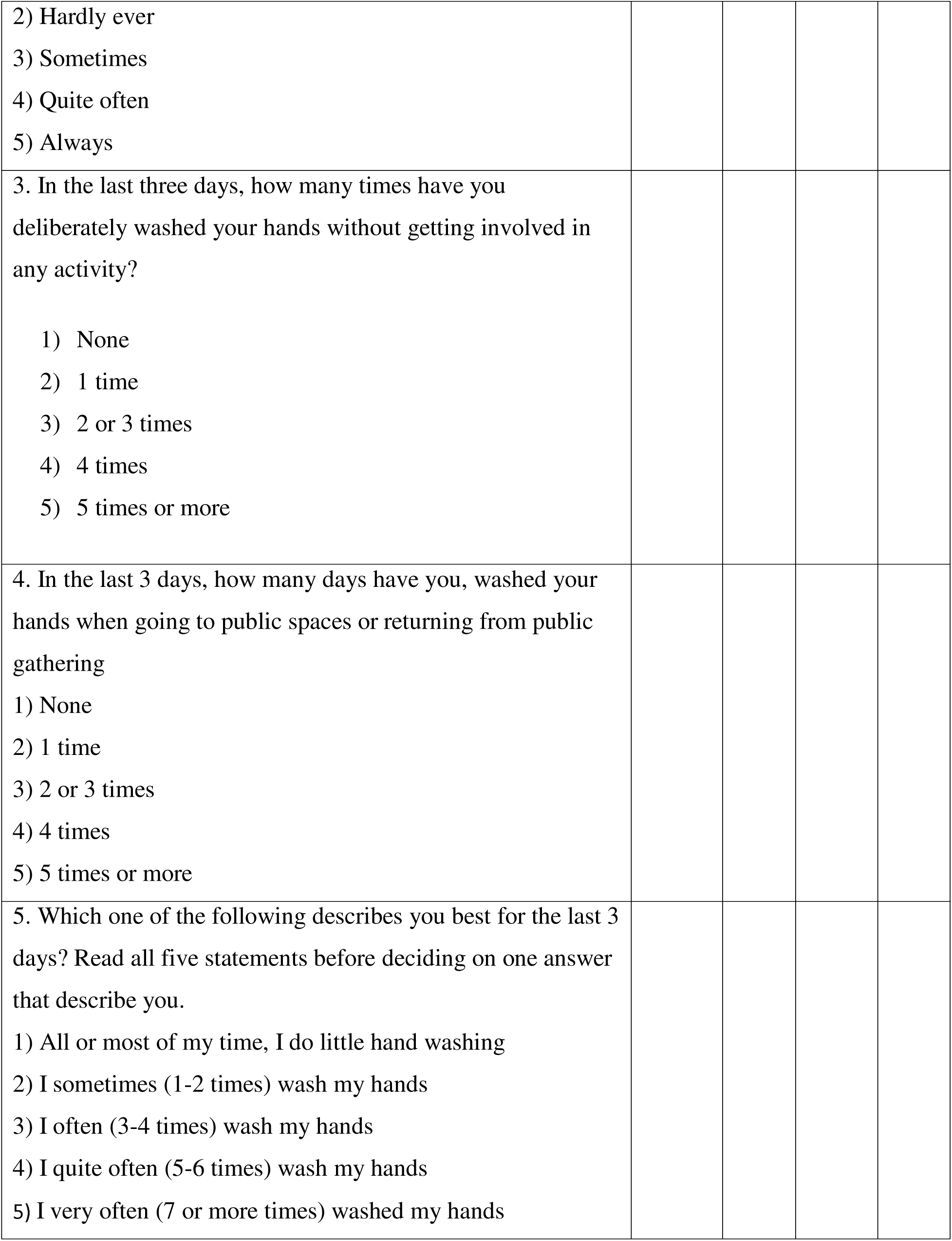

### FINAL DRAFT-THE HANDWASHING INDEX (HWI)

1. How many times, have you been hand washing, in the past **three (3)** days?

1. None
2. 1-2 times
3. 3-4 times
4. 5-6 times
5. 7 times or more
2. How often are you active at washing your hands in the last **3 days**?

1. I don’t do any
2. Hardly ever
3. Sometimes
4. Quite often
5. Always
3. In the last 3 days, how many days have you, washed your hands when going to public spaces or returning from public gathering

1. None
2. 1 time
3. 2 or 3 times
4. 4 times
5. 5 times or more
4. Which one of the following describes you best for the last 3 days? Read all five statements before deciding on one answer that describe you.

1. All or most of my time, I do little hand washing
2. I sometimes (1-2 times) wash my hands
3. I often (3-4 times) wash my hands
4. I quite often (5-6 times) wash my hands
5. I very often (7 or more times) washed my hands

**[Number of Items-4]**

#### INSTRUCTIONS

Scores of 1 indicate low activity, and 5 indicate high activity. Taking the mean of all activity of each item gives a composite or average score. The categorization of average scores is as follows:

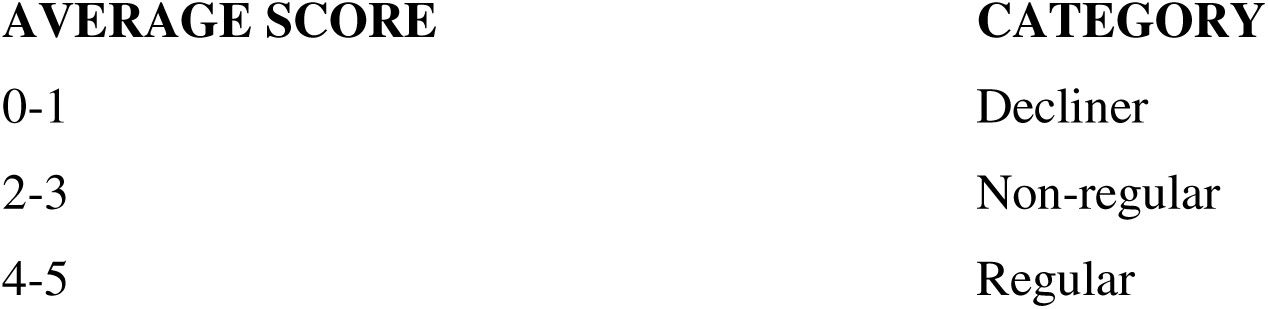

